# Mapping the human helminthiases: a systematic review of geospatial tools in medical parasitology

**DOI:** 10.1101/2020.10.30.20223529

**Authors:** Catherine G. Schluth, Claire J. Standley, Shweta Bansal, Colin J. Carlson

**Author notes:** Correspondence should be directed to.

## Abstract

Helminthiases are a class of neglected tropical diseases that affect at least one billion people worldwide, with a disproportionate impact in resource-poor areas with limited disease surveillance. Geospatial methods can offer valuable insights into the burden of these infections, particularly given that many are subject to strong ecological influences on the environmental, vector-borne, or zoonotic stages of their life cycle. In this study, we screened 6,829 abstracts and analyzed 485 studies that use maps to document, infer, or predict transmission patterns for over 200 species of parasitic worm. We found that quantitative mapping methods are increasingly used in medical parasitology, drawing on One Health surveillance data from the community scale to model geographic distributions and burdens up to the regional or global scale. However, we found that the vast majority of the human helminthiases may be entirely unmapped, with research effort focused disproportionately on a half-dozen infections that are targeted by mass drug administration programs. Entire regions were also surprisingly under-represented in the literature, particularly southern Asia and the Neotropics. We conclude by proposing a shortlist of possible priorities for future research, including several neglected helminthiases with a burden that may be substantially underestimated.

## Introduction

Infections with parasitic worms, or *helminthiases*, have a massive global burden on human health. More than a billion people are infected with soil-transmitted helminths alone (1), with a total burden of over 3.3 million disability-adjusted life years (DALYs) (2); these may be underestimates, given more recent estimates that hookworm alone may account for more than 4 million DALYs (3). The burden of these infections is highly heterogeneous over space, from case clustering at the community level up to the global scale. Helminthiases persist and recur disproportionately where healthcare systems are too limited for routine treatment, preventative therapies, and case management; but these areas are, conversely, often the places where disease surveillance is most limited, and so the burden of these infections is most poorly characterized.

Many previous studies have therefore identified geospatial analysis as a key part of scientific and clinical work on helminthiases. As a basic tool of descriptive epidemiology, maps are one of the simplest, easiest way to visualize data, communicate risk, and engage local communities in participatory research methods. Morever, geospatial modeling can help fill knowledge gaps about the prevalence or incidence of infectious diseases in under-sampled regions, turning clinical data into a more continuous view of transmission. With enough data, this approach can be used to translate local prevalence surveys into regional and global estimates of incidence or burden. Along the way, geospatial modeling often illuminates environmental and social risk factors for disease, and perhaps most importantly, helps practitioners target, evaluate, and improve interventions.

Previous reviews of infectious disease cartography have evaluated research effort and set priorities for future pathogens to map (4; 5), but have only minimally addressed the human helminthiases. A systematic analysis of research trends could help identify the limitations of existing data, target interventions more effectively, and broaden the scope of helminth research and control. Here, out of over 6,000 candidate mapping studies, we examine a total of 485 scientific studies that developed empirical maps of over 200 helminth species known to infect humans. From these studies, we evaluate trends in research methodology and scope, highlight global gaps in research effort, and propose a list of neglected helminthiases that researchers (and surveillance systems) could prioritize in future geospatial studies.

## Methods

### Identifying candidate species

To compile a list of human-associated helminth species, we used a recently-published dataset of host-parasite associations curated by the Natural History Museum in London (NHM). (6) From these data, we compiled a list of human helminthiases by searching for associations with *Homo sapiens*, and recording the number of references listed for each species. There were 407 helminth species on this initial list. To verify whether each helminth species is still considered taxonomically valid and is capable of infecting humans, we manually searched for records of human infection for each species. In Google Scholar, we used the search queries “[species name]” and “human*” to search for records of human infection. In Google, we used the search queries “[species name]” and “syn*” to determine if species with no records or only old records of human infection have since been renamed. We removed a species from the study if we could find no evidence that the species infects humans, the species name was found to be synonymous with a more recent species name on the list, there was conflicting evidence as to whether the species can infect humans, or the species was found to infect humans only in a hybrid with another helminth species. This left a total of 232 taxonomically-valid human helminthiases.

### Systematic review

Taking an iterative approach to data collection, we used *Wuchereria bancrofti* as a test species to determine which search terms would be most effective to retrieve helminth mapping studies. On Google Scholar, we used the search queries “*Wuchereria bancrofti*” and “mapping.” Based on an informal analysis of the results, we selected a final set of keywords that consistently signaled that empirical spatial analysis was undertaken. Our final search query was “[species name]” and (“SaTScan” or “MaxEnt” or “spatial cluster*” or “spatial analysis” or “geospatial” or “ecological niche model*” or “mapping” or “nearest neighbor” or “spatial GLM*”). To identify candidate studies, we searched PMC and PubMed for these terms with each of the 232 helminth species, one by one. Our search may have missed mapping studies that were written and published in other languages, as well as grey literature produced by health ministries or non-governmental organizations; as such, our study represents only a snapshot of the retrievable literature.

Our literature review was conducted between November 2018 and May 2019 (and is, as such, limited to the pre-pandemic period), following PRISMA guidelines (see Figure S1). We included studies in our dataset if they presented novel data or a new modeling product representing the known or predicted spatial distribution of a helminth species, the condition(s) it causes, and/or the medication used to treat it; studies were excluded if they did not use empirical data to generate either a map-based visualization or a spatial model of human helminthiasis infection over space. Two authors conducted the review and both verified each study that was selected for inclusion. In total, we found 485 studies that mapped a total of 45 helminth species. For each study in the final dataset and analysis, we recorded all available information on: the full binomial nomenclature of helminth species being mapped; the citation for and link to each study; the year each study was conducted; the spatial scope of each study; the specific methodologies used in each mapping effort; the sample size and type in each study; whether each study addressed uncertainty and population at risk; whether each study examined coendemicity and/or coinfection among helminth species or between helminth species and other diseases; and whether each study had its data publicly archived.

### Ontology of study methodology

We classified the mapping methodology of studies into a handful of non-exclusive categories:

- **Grey data** describes the presentation of spatial point data of either cases or positive-negative testing results. For attempts to develop *post hoc* databases and risk maps, these are data that could be heads-up digitized and reused (provided they have not been jittered for data security and anonymization).
- **Prevalence mapping** refers to raw or aggregated prevalence data presented on a map; like grey data, this is a presentation of raw data, but with more granularity with respect to intensity of transmission. For our purposes, this also includes non-prevalence quantitative measures of transmission intensity, like fecal egg counts.
- **Prevalence modeling** refers to using statistical models to analyze or reconstruct patterns of prevalence, involving either a model with an explicit spatial component or spatial covariates (like climate data), or spatial autocorrelation analyses such as Moran’s I or autocorrelograms. (For example, a binomial logistic regression that is stratified by age and sex alone would not qualify for this; such a model incorporating distance to rivers, gridded rainfall, or a conditionally autoregressive term would qualify.)
- **Cluster analysis** refers to spatial clustering methods like SaTSCAN or directional distribution models (standard deviation ellipse), a subset of prevalence modeling focused on identifying specific, discrete points and areas of case clusters or transmission hotspots.
- **Risk mapping** refers broadly to the projection of modeled risk surfaces over a continuous area or at regional levels; this involves at least some amount of inference of risk, from a model of prevalence or occurrence, and visual presentation of *modeled* results (i.e., not hand-drawn or expert-informed maps).
- **Ecological niche modeling** is a subtype of risk mapping, for when authors explicitly refer to habitat suitability models, ecological niche models, or species distribution models as the methodology being used. Risk maps with ecological covariates are arguably habitat suitability models *sensu lato*, but not necessarily part of that specific body of literature.
- Finally, **endemicity mapping** refers to the delineation of known or suspected zones of endemicity or transmission, based on historical or published data reused or consolidated to identify likely zones. For example, this can be the identification of possibly at-risk communities, or the mapping of survey results at the national level for a whole continent.

## Results

### Helminthiases are a topic of growing interest in medical geography

We found a total of 485 studies that mapped human helminthiases across a mix of scales, regions, pathogens, and purposes. The number of helminth mapping studies has steadily increased since the year 2000 (Figure 1), and the field is still growing rapidly. Across all time periods, we found that most studies use maps first and foremost as a data visualization tool (case occurrence data or prevalence maps; Figure 2); however, the last decade has seen a particular shift towards advanced statistical modeling and machine learning approaches. In particular, tools from ecological niche modeling began to be used around 2007 to 2010, when the most popular algorithms (MaxEnt and GARP) began to cross over into medical geography. This particular approach to predictive modeling is continuing to become more popular as more disease ecologists become involved in neglected tropical disease research. In the last decade, we also observed a methodological shift away from studies using licensed software like ArcGIS, and increasingly taking advantage of open-source software like QGIS and GRASS, or console programs like R and Python. These accessible softwares can be easily used by researchers and stakeholders without the financial barriers of proprietary softare that are prohibitive even for many in the Global North.

**Figure 1:**
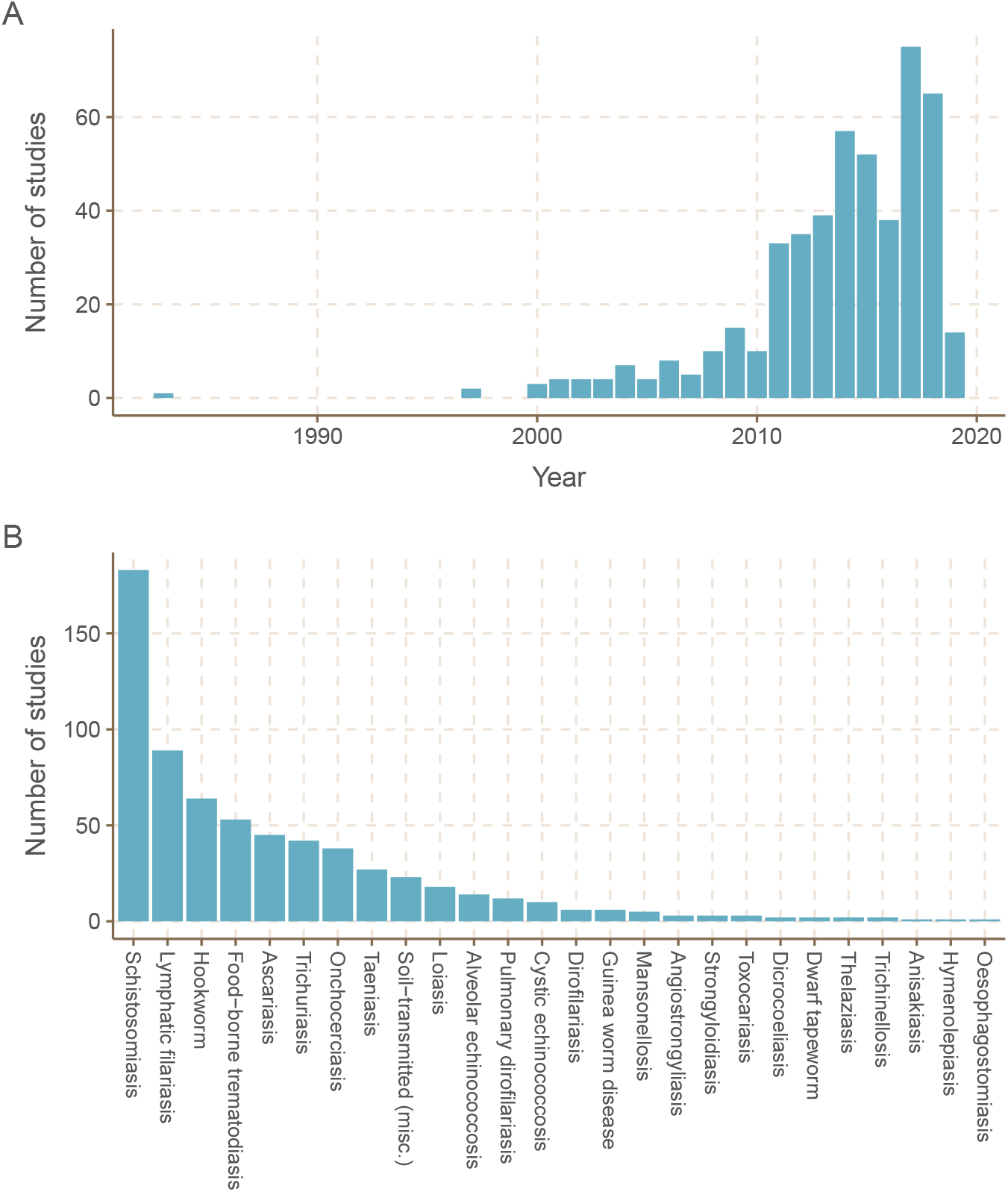
(A) Efforts to map the human helminthiases have increased over time. (B) Spatial data for a few helminthiases makes up the majority of all human helminth spatial data. The 45 helminth species with spatial data were grouped together by the conditions they cause (e.g. *Wuchereria bancrofti* and *Brugia malayi* grouped as lymphatic filariasis).

**Figure 2:**
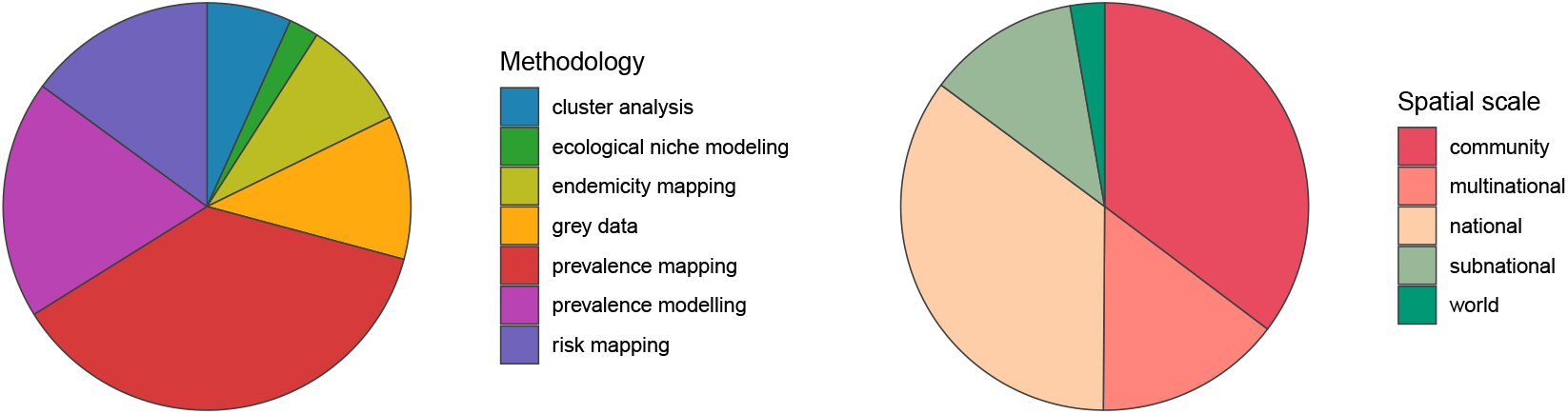
Existing human helminth spatial data predominantly comes from small-scale prevalence mapping studies. Studies containing spatial data on human helminthiases were characterized by spatial scale and methodology, with several studies employing more than one methodology.

### Spatial scale and global gaps

We found that the vast majority of mapping studies were boots-on-the-ground epidemiological research conducted in a single community or a handful of communities within a single country (Figure 2). We found relatively few large-scale (multinational to global) prevalence or risk maps, likely because the raw prevalence data are unsynthesized in a modelable format or, for many infections, have never been collected at sufficient scale. We also found that the distribution of research effort has been strikingly uneven (Figure 3). Hotspots of research in China, Brazil, and tropical Sub-Saharan Africa reflect a mix of population size, infectious disease burden, and unique aspects of the medical parasitology community of practice (e.g., Fiocruz in Brazil). However, we found major research gaps in South and Southeast Asia, the Middle East, and Latin America and the Caribbean, despite the high parasite burden faced by many communities in these regions (e.g., (7)). Overall, these findings suggest that the global burden of many helminthiases might be underestimated, especially if parasite prevalence is high in research and surveillance coldpots.

**Figure 3:**
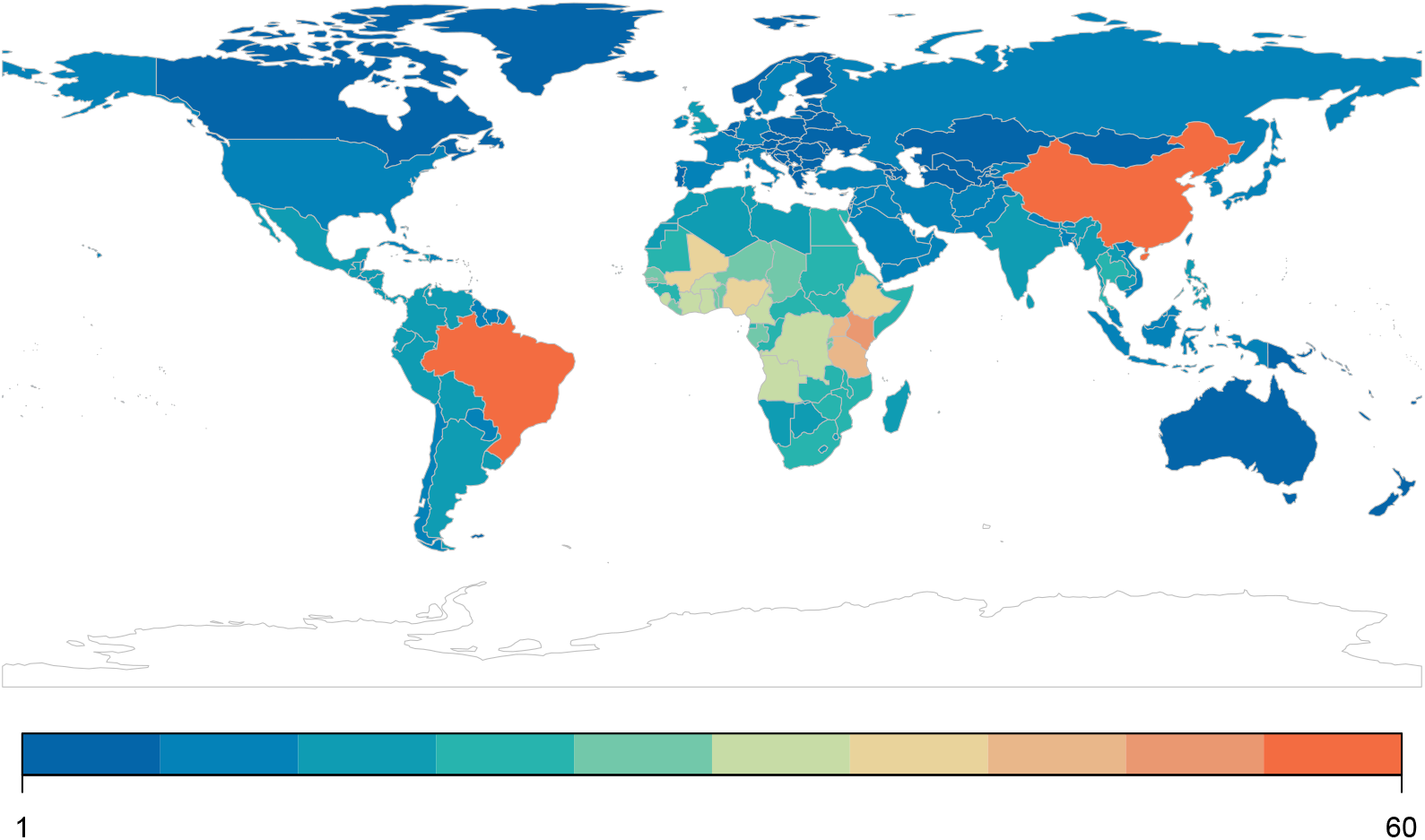
Most published spatial research on human helminthiases describes incidence and burden in Sub-Saharan Africa, China, and Brazil.

### Most human helminthiases are unmapped or undermapped

Of the 232 species targeted by our systematic review, only 45 had any associated studies— indicating nearly 200 unmapped species of human parasites. Out of those 45, a half-dozen of the best-known conditions—the major vector-borne helminthiases (schistosomiasis, lymphatic filariasis, and onchocerciasis) and soil-transmitted helminthiases (hookworm, *Ascaris*, and *Trichuris*)— account for the vast majority of research effort (Figure 1). Unsurprisingly, these infections account for the majority of the total burden of helminthiases on global health and poverty. Ascariasis is the most common helminth infection in the world, thought to infect between 737 million and 872 million people worldwide (8), and is a major cause of stunting and malnutrition in children (9). *Trichuris* and hookworm have a similarly massive burden, infecting roughly 435 million and 450 million people worldwide, respectively (8). Schistosomiasis infects between 179 million and 200 million people worldwide (8), with a high burden especially in HIV-coendemic areas. Aside from these infections, all other human helminthiases are generally presumed to infect fewer than 100 million people worldwide.

### Feedbacks between mapping and interventions

Research effort also reflects feedbacks among mapping work, ease of treatment, and scale of interventions. A small number of infections are targeted by MDA programs, both because of cheap widely-available treatments and because they account for the highest global burden. These programs are naturally complimentary with spatial analysis: defining the boundaries of a community, testing people or animals for helminthiases, and updating endemicity maps is one of the easiest ways to visualize burden and decide on the frequency and distribution of drug administration. This ongoing feedback of prevalence studies, GIS work, and targeted drug administration has been a key part of successful MDA efforts over the past 20 years, not just to tailor efforts but also to measure their success and justify ongoing funding. These programs are therefore the main reason that broad, synthetic data and cartography are possible for a small subset of the best-mapped helminthiases (i.e., soil-transmitted helminths, schistosomiasis, lymphatic filariasis, and onchocerciasis). Conversely, we found that most infections without readily available antihelminthic treatments were relatively understudied, or never appeared in our dataset.

Soil-transmitted helminths are also exceptional in that most have a relatively simple life cycle; as such, prevalence data in humans are usually likely to capture the extent of transmission, (though see (10)). In contrast, many under-represented helminth species were zoonotic, likely because complex life cycles or wildlife hosts make them more challenging targets for surveillance. When non-human hosts are studied, they are almost always livestock (especially cattle and sheep), pets (cats and dogs), or synanthropic wildlife (rats and mice); true wildlife hosts account for a small fraction of studies (Figure 4). Some of the most understudied helminthiases are the ones that complete parts of their life cycle in hosts that are particularly difficult to sample, like fish and marine mammals (e.g., *Anisakis* or *Diphyllobothrium*). Similarly, vector-borne transmission adds another layer of ecological influence, which can make ecological models more useful, but primary data collection more challenging; we found more studies collected spatial data on snails (the “vectors,” or more accurately intermediate hosts, of schistosomiasis) than on more mobile vectors like mosquitoes or flies. Surprisingly, even for soil-transmitted species, we found that environmental sampling is nearly never reported (Figure S2).

**Figure 4:**
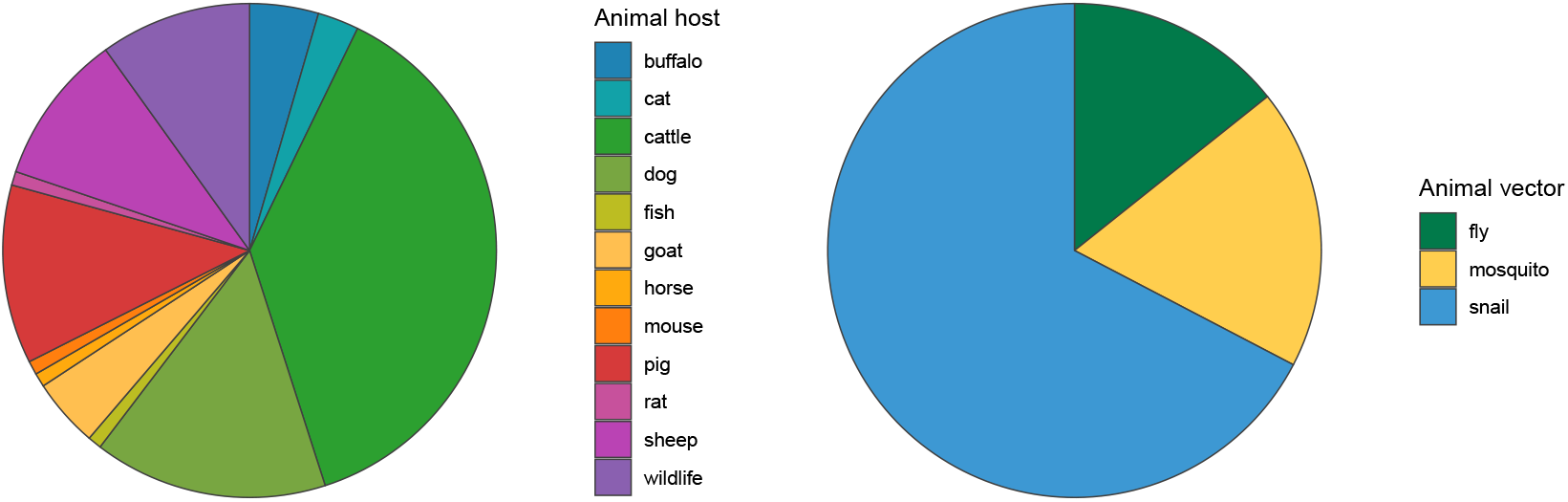
Among studies that map other helminth hosts or helminth vectors, studies mapping less mobile hosts and vectors predominate. Baboons, antelope, and wild boar were classified as wildlife hosts; some studies mapped multiple non-human hosts

### Coinfections, coendemicity, and syndemic interactions

We found that a surprising number of mapping studies directly addressed helminth coinfections and coendemicity. Many surveillance programs—especially those guiding mass drug administration— inherently collect data on multiple helminthiases at once (e.g., Kato-Katz screening can detect *Ascaris, Trichuris*, schistosomiasis, hookworm, and other eggs of other parasites as well). Studies that map hotspots of coinfection (e.g., (11; 12; 13)) can help prioritize where treatment might have the greatest social and economic benefit. These approaches can also address more complicated syndemic interactions (14): for example, onchocerciasis control programs often use ivermectin, a drug that can cause severe neurological complications or even death when administered to a patient with loiasis (15). Increasingly, mapping studies have been used to address *Onchocerca*-*Loa* coendemicity in west and central Africa, helping practitioners to delineate where ivermectin can be administered safely, and where other interventions like vector control might be safer and more effective.

Finally, we found over a dozen studies that addressed coinfections or coendemicity of helminthiases with other infections. Most of these studies focused on malaria: three studies mapped coinfections with hookworm (13; 16; 17), two with schistosomiasis (18; 19), and another with lymphatic filariasis (20). Integrated mapping can address different aspects of syndemic interactions: for example, some helminthiases share a preventable transmission route with other pathogens (e.g., *Anopheles* mosquitoes transmit both malaria and lymphatic filariasis); others are treatable with the same drugs (artemisin, a widely-used antimalarial, also targets immature schistosomes (21); recent evidence suggests ivermectin in bloodmeals may reduce *Anopheles* mosquito lifespan (22; 23)). Perhaps the most elusive facet of helminthiases’ burden is their immunomodulatory effects, which can have unpredictable impacts on other diseases: for example, while *Schistosoma mansoni* or hookworm infections may increase susceptibility to malaria, *S. haematobium* infections may confer protection against severe malaria (24; 25). In any of these contexts, helminthiases are worth considering in broader efforts to measure and reduce the global burden of disease.

## Discussion

Here, we screened over 6,000 studies, and found extensive literature on the human helminthiases that incorporated geospatial approaches (nearly 500 studies). However, we found that most of these studies were focused on a half-dozen or so parasites with a simple life cycle, available low-cost treatments, and the greatest global burden—the circumstances that make elimination programs a cost-effective investment. For the vast majority of human helminthiases, we found no geospatial data or analysis of any kind in the studies we reviewed. Some of these parasites may only sporadically infect humans, but several others are known to have an uncertain but likely medium-to-high global burden. Often, these neglected helminthiases have a complex (zoonotic or vector-borne) life cycle that both complicates surveillance and limits the feasibility of vertical control programs (especially if elimination is precluded by non-human reservoirs).

For these neglected infections, there are many opportunities for mapping work to both establish a clearer baseline on global burden, and to support One Health interventions that include vector control, community sanitation, food safety, livestock vaccination, routine deworming of household pets, and similar practices. In service of this goal, we propose a shortlist of several notable but neglected human helminthiases that were underrepresented in the literature (Box 1).

### Box 1. An (incomplete) list of high-priority helminthiases for geospatial research

1. Angiostrongyliasis (*Angiostrongylus cantonensis*)
2. Hepatic and intestinal capillariasis (*Capillaria hepatica, Capillaria philippinensis*)
3. Carcinogenic food-borne trematodiases (*Clonorchis sinensis, Opisthorchis viverrini*)
4. Guinea worm disease* (*Dracunculus medinensis*) (*see text)
5. Echinococcosis (*Echinococcus granulosus, E. multilocularis*)
6. Gastrodiscoidiasis (*Gastrodiscoides hominis*)
7. Dwarf tapeworm (*Hymenolepis nana*)
8. Mansonellosis (*Mansonella perstans, M. ozzardi, M. streptocerca*)
9. Strongyloidiasis (*Strongyloides stercoralis*)
10. Taeniosis and cysticercosis (*Taenia solium*)

### Opportunities for global burden (re-)estimation

Several helminthiases with a global distribution have a likely-high but uncertain burden, which could be clarified by a coordinated data synthesis and geospatial modeling effort—potentially motivating more global investment in prevention and treatment. For example:

#### Echinococcosis

*Echinococcus granulosus* and *E. multilocularis* are zoonotic tapeworms that cause cystic and alveolar echinococcosis, respectively. These infections typically remain asymptomatic for years until cysts grow large enough to disrupt organ function; when they rupture, or (in the latter case) result in liver failure, case fatality rates are relatively high (26). Between the two infections, recent estimates place the global burden around roughly 19,000 deaths per year out of 1 million active cases (27). Treatment is difficult, and may require surgery, but infections can also be prevented with a One Health strategy that includes slaughterhouse hygiene and deworming dogs. Global summaries of prevalence data at national or subnational levels were recently compiled not just for human hosts, but also wildlife and domesticated hosts (28); these data could be readily applied to more detailed, fine-scale geospatial modeling.

#### Taeniasis and cysticercosis

A zoonotic parasite of swine, *Taenia solium* is endemic worldwide in communities with poor sanitation and consumption of undercooked pork. Intestinal infection with the adult tapeworm is usually mild, but fecal-oral transmission between humans leads to paratenic infections that can form on the brain or on the spinal cord. These severe infections cause at least 28,000 deaths annually (27), and account for at least a third of all epilepsy cases in endemic areas (29). Estimates range between 2-6 million infections worldwide (8; 27), but regional estimates (e.g. 1.2 million attributable epilepsy cases in India alone (30)) suggest these are underestimates. A high-resolution global estimate of burden could help target One Health interventions pairing MDA for taeniosis with pig vaccination, which can eliminate the pathogen over just 4-5 years (31).

#### Strongyloidiasis

*Strongyloides stercoralis* is a soil-transmitted nematode that infects at least 30-100 million people in rural communities without proper sanitation (32). Strongyloidiasis is often asymptomatic, but can be life-threatening in immunocompromised individuals (33). Our systematic review identified just three efforts to map this parasite—all national or community studies—highlighting an opportunity to consolidate existing surveillance data, and develop high-resolution maps of endemicity and burden. One recent study takes an important step towards filling this gap by developing a global ecological niche model for strongyloidiasis (34), but data remain limited and more systematic efforts are needed.

#### Dwarf tapeworm

*Hymenolepis nana* is one of the most common cestode parasites of humans. These infections are generally asymptomatic in adults, but more severe in children, especially when comorbid with malnutrition (35). Estimates of regional *H. nana* prevalence vary substantially, ranging from 0.2 to 28.4% in Asia and from 0.9 to 23% in the Americas (36). Our literature review found only two mapping studies on *H. nana*, both community-based studies in Angola and Ghana (16; 37). Future work could consolidate fine-scale surveillance datasets, and align them with other geospatial research on malnutrition and stunting (e.g., (38)).

### Opportunities for global distribution or risk mapping

For several other helminthiases with a global distribution, a baseline global map of endemicity—or a risk map of the zoonotic niche and the socioenvironmental risk factors for infection—might be a substantial step forward. For example:

#### Angiostrongyliasis

More commonly called rat lungworm, *Angiostrongylus cantonensis* is a rare but emerging infection endemic to Asia and the Pacific that causes eosinophilic meningitis Some infections are self-limiting, but others may cause significant neurological damage or death. The geographic range of the parasite has expanded over time, potentially facilitated by climate change and human movement, with several thousand cases reported worldwide (39; 40). Existing literature on the parasite’s global distribution has worked from sparse data (40; 41), and future work could more directly address the parasite’s zoonotic niche in both intermediate hosts (snails and slugs) and the ultimate host (rats).

#### Mansonellosis

An infection that has been called “the most neglected human filariasis” (42), *Mansonella perstans* alone is thought to infect over 100 million people (43), yet there are currently no large-scale control programs targeting any *Mansonella* species. Our literature review identified nine studies mapping mansonellosis, including a map of global endemicity. Future research could aim to generate a high-resolution global risk map to guide vector control efforts, particularly given that its *Culicoides* midge vectors also transmit several emerging infections (including bluetongue virus, Oropouche virus, and African horse sickness).

#### Guinea worm disease

Unlike other infections on our shortlist, Guinea worm disease cannot be considered neglected. Decades of control efforts brought *Dracunculus medinensis* close to being the first globally-eradicated parasite, though canine reservoirs now jeopardize that progress (44; 45). Despite the small number of remaining transmission foci, Guinea worm was once found throughout the tropics. Little geospatial data has been consolidated on this century-long range contraction; the best available maps of its original range are hand-drawn estimates from the 1950s (46). Remapping the historical distribution of *D. medinensis* with modern technology and modeling methods could offer some insights into the previous successes of eradication campaigns, clarify the ongoing role of zoonotic reservoirs (47), and offer fresh motivation to eliminate Guinea worm disease on the last remaining continent.

### Opportunities for regional and community-based mapping

For a handful of the rarest or most neglected helminthiases, more boots-on-the-ground studies of parasite prevalence in local communities are still needed, forming the quantitative basis of broader estimates of parasite distribution and prevalence.

#### Carcinogenic food-borne trematodiases

*Opisthorchis viverrini* and *Clonorchis sinensis* are trematode parasites of fish (48), both of which seriously increase the risk of bile duct cancer (cholangiocarcinoma). (48) The burden of *O. viverrini* is localized to southeast Asia and particularly Thailand, which has the highest incidence of cholangiocarcinoma worldwide (49); opisthorchiasis prevalence reaches 70% in some communities, costing the country an estimated $120 million USD annually (48). *C. sinensis* has a much wider range throughout east Asia, infecting an estimated 35 million people (50). Our systematic review found no community-based mapping efforts for *C. sinensis*, and very few for *O. viverrini*.

#### Capillariasis

*Capillaria hepatica* and *C. philippinensis* cause hepatic and intestinal capillariasis, respectively. The former is believed to only infect humans rarely, while outbreaks of the latter have been sizable; both can be deadly, and are challenging to diagnose (51). Human infections with both parasites are apparently rare but widespread: *C. philippinensis* can be found throughout Asia and parts of North Africa and the Middle East, and *C. hepatica* is found in wildlife worldwide (51; 52). However, our literature search did not identify any efforts to map either species.

#### Gastrodiscoidiasis

*Gastrodiscoides hominis* is a poorly characterized food-borne zoonotic fluke (53), capable of causing diarrhea and malnutrition (54; 55), and even death (54). *G. hominis* is thought to be highly prevalent in India, but cases have been observed throughout Asia and parts of Africa (54). Our literature search did not identify any efforts to map *G. hominis* infection.

### Broader opportunities for geospatial research

Infectious diseases are rarely a stationary target for mapping and modeling. While geospatial methods are already widely used to track the progress of elimination programs, we finally note that these tools will also have value as climate change drives shifts in the geographic distribution and burden of helminthiases, their vectors, and their wildlife reservoirs. As temperatures rise, the range of hookworm is expected to expand toward the southernmost regions of Africa (56); as *Anopheles* mosquitoes and other vectors undergo geographic range shifts (57), they may introduce parasites into new regions or undo progress towards elimination. In other cases, climate change may make environmental conditions less hospitable for parasites (58): angiostrongyliasis may lose range as China warms (59), and the snail intermediate hosts of schistosomiasis may begin to shrink as their habitats grow hotter and more arid (56). Ecological modeling will help triage the greatest climate-related risks to human health, while other mapping methods will be critical to document the impacts of climate change in real-time. In that light, One Health data on parasitic infections in humans, livestock, domestic animals, and wildlife all become even more valuable—particularly when shared openly with sufficient geospatial metadata for reuse.

## Data Availability

Data will be made available upon publication

## Declarations

### Availability of data and material

All data will be made available on Zenodo at the time of publication.

### Competing interests

The authors have no competing interests to declare.

### Funding

The authors have no funding to declare.

### Authors’ contributions

All authors contributed to the writing and conception of the manuscript.

## Acknowledgements

The authors thank the relentless work of the scientists, clinicians, and volunteers who have generated the scientific literature reviewed in this study.

## Supporting Information

**Supplementary Figure 1:**
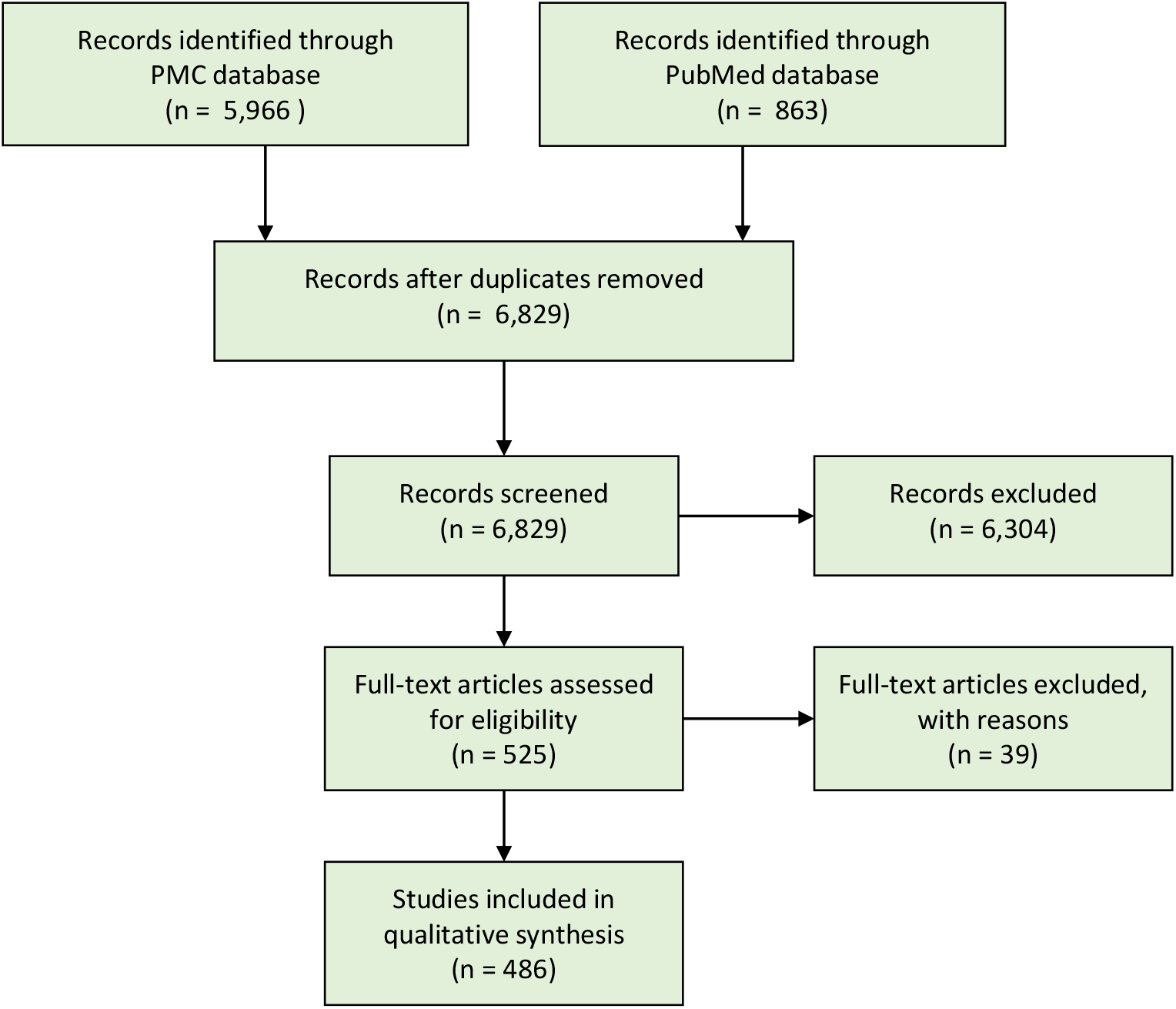
Systematic review procedure, following PRISMA reporting guidelines.

**Supplementary Figure 2:**
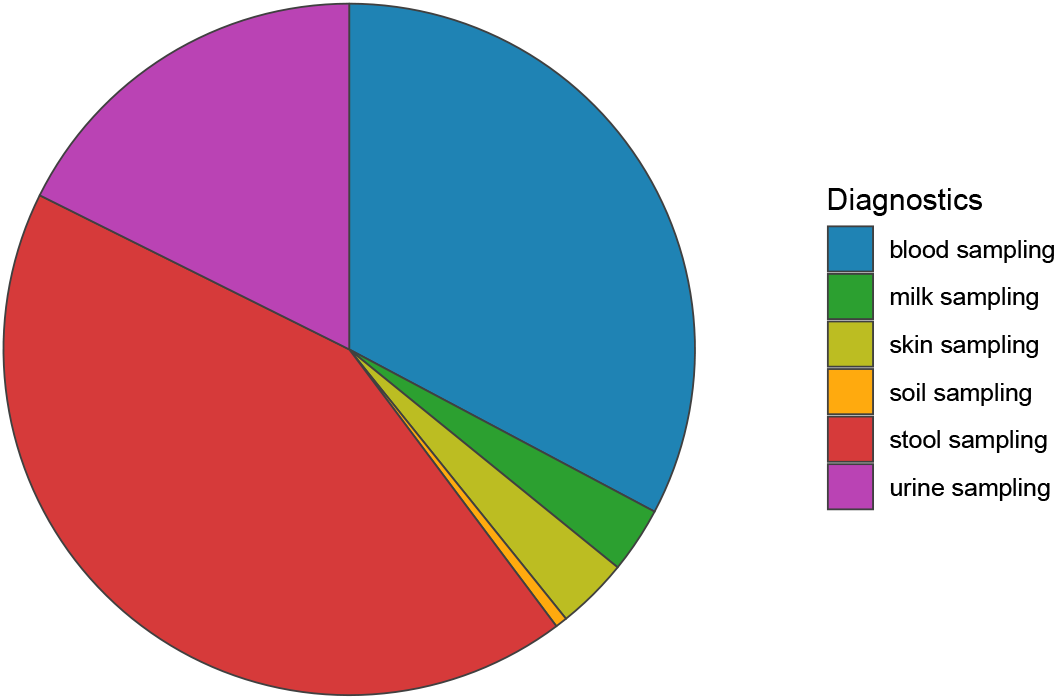
Diagnostic method used to collect primary data in 357 studies that report new data from field surveillance.

## Notes

### Competing Interest Statement

The authors have declared no competing interest.

### Funding Statement

No external funding to declare

### Summary of Updates

Small reformatting and some paring down of text

## References

[1] GBD 2016 DALYs and HALE collaborators. Global, regional, and national disability-adjusted life-years (DALYs) for 333 diseases and injuries and healthy life expectancy (HALE) for 195 countries and territories, 1990–2016: a systematic analysis for the Global Burden of Disease Study 2016. The Lancet. 2017;390(10100):1260–1344.

[2] Pullan RL, Smith JL, Jasrasaria R, Brooker SJ. Global numbers of infection and disease burden of soil transmitted helminth infections in 2010. Parasites and Vectors. 2014;7(37).

[3] Weatherhead JE, Hotez PJ, Mejio R. The global state of helminth control and elimination in children. Pediatric Clinics of North America. 2017;64(4).

[4] Hay SI, Battle KE, Pigott DM, Smith DL, Moyes CL, Bhatt S, et al. Global mapping of infectious disease. Philosophical Transactions of the Royal Society B: Biological Sciences. 2013;368(1614):20120250.

[5] Pigott DM, Howes RE, Wiebe A, Battle KE, Golding N, Gething PW, et al. Prioritising infectious disease mapping. PLoS Neglected Tropical Diseases. 2015;9(6):e0003756.

[6] Dallas T. helminthR: an R interface to the London Natural History Museum’s host–parasite database. Ecography. 2016;39(4):391–393.

[7] Salam N, Azam S. Prevalence and distribution of soil-transmitted helminth infections in India. BMC Public Health. 2017;17(201).

[8] GBD 2016 Disease and Injury Incidence and Prevalence Collaborators. Global, regional, and national incidence, prevalence, and years lived with disability for 328 diseases and injuries for 195 countries, 1990–2016: a systematic analysis for the Global Burden of Disease Study 2016. The Lancet. 2017;390(10100):1211–1259.

[9] World Health Organization. Helminth control in school[U+2010]age children: a guide for managers of control programmes. WHO Technical Report Series. 2011;2:1–90.

[10] Nejsum P, Betson M, Bendall R, Thamsborg SM, Stothard J. Assessing the zoonotic potential of Ascaris suum and Trichuris suis: looking to the future from an analysis of the past. Journal of Helminthology. 2012;86(2):148–155.

[11] Raso G, Vounatsou P, Singer BH, N’Goran EK, Tanner M, Utzinger J. An integrated approach for risk profiling and spatial prediction of Schistosoma mansoni–hookworm coinfection. Proceedings of the National Academy of Sciences of the United States of America. 2006;103(18):6934–6939.

[12] Pullan RL, Bethony JM, Geiger SM, Cundill B, Correa-Oliveira R, Quinnell RJ, et al. Human Helminth Co-Infection: Analysis of Spatial Patterns and Risk Factors in a Brazilian Community. PLoS Neglected Tropical Diseases. 2008;2(12).

[13] Brooker S, Clements AC. Spatial heterogeneity of parasite co-infection: Determinants and geostatistical prediction at regional scales. International Journal for Parasitology. 2009;39(5):591–597.

[14] Singer M, Bulled N, Ostrach B, Mendenhall E. Syndemics and the biosocial conception of health. The Lancet. 2017;389(10072):941–950.

[15] Wanji S, Ndongmo WP, Fombad FF, Kengne-Ouafo JA, Njouendou AJ, Tchounkeu YF, et al. Impact of repeated annual community directed treatment with ivermectin on loiasis parasitological indicators in Cameroon: Implications for onchocerciasis and lymphatic filariasis elimination in areas co-endemic with Loa loa in Africa. PLoS Neglected Tropical Diseases. 2018;12(9).

[16] Adu-Gyasi D, Asante KP, Frempong MT, Gyasi DK, Iddrisu LF, Ankrah L, et al. Epidemiology of soil transmitted Helminth infections in the middle-belt of Ghana, Africa. Parasite Epidemiology and Control. 2018;3(3):e00071.

[17] Brooker SJ, Pullan RL, Gitonga CW, Ashton RA, Kolaczinski JH, Kabatereine NB, et al. Plasmodium–helminth coinfection and its sources of heterogeneity across east Africa. Journal of Infectious Diseases. 2012;205(5):841–852.

[18] Kabatereine NB, Standley CJ, Sousa-Figueiredo JC, Fleming FM, Stothard JR, Talisuna A, et al. Integrated prevalence mapping of schistosomiasis, soil-transmitted helminthiasis and malaria in lakeside and island communities in Lake Victoria, Uganda. Parasites and Vectors. 2011;4(1):232.

[19] Doumbo S, Tran TM, Sangala J, Li S, Doumtabe D, Kone Y, et al. Co-infection of long-term carriers of Plasmodium falciparum with Schistosoma haematobium enhances protection from febrile malaria: a prospective cohort study in Mali. PLoS Neglected Tropical Diseases. 2014;8(9):e3154.

[20] Stensgaard AS, Vounatsou P, Onapa AW, Simonsen PE, Pedersen EM, Rahbek C, et al. Bayesian geostatistical modelling of malaria and lymphatic filariasis infections in Uganda: predictors of risk and geographical patterns of co-endemicity. Malaria Journal. 2011;10(1):298.

[21] Bergquist R, Elmorshedy H. Artemether and Praziquantel: Origin, Mode of Action, Impact, and Suggested Application for Effective Control of Human Schistosomiasis. Tropical Medicine and Infectious Disease. 2018;3(4).

[22] Derua YA, Kisinza WN, Simonsen PE. Differential effect of human ivermectin treatment on blood feeding Anopheles gambiae and Culex quinquefasciatus. Parasites and Vectors. 2015;8.

[23] Mekuriaw W, Balkew M, Messenger LA, Yewhalaw D, Woyessa A, Massebo F. The effect of ivermectin® on fertility, fecundity and mortality of Anopheles arabiensis fed on treated men in Ethiopia. Malaria Journal. 2019;18(357).

[24] Donahue RE, Cross ZK, Michael E. The extent, nature, and pathogenic consequences of helminth polyparasitism in humans: A meta-analysis. PLoS Neglected Tropical Diseases. 2019;13(6).

[25] Adegnika AA, Kremsner PG. Epidemiology of malaria and helminth interaction: a review from 2001 to 2011. Current Opinion in HIV and AIDS. 2012;7(3).

[26] Wen H, Vuitton L, Tuxun T, Li J, Vuitton DA, Zhang W, et al. Echinococcosis: Advances in the 21st Century. Clinical Microbiology Reviews. 2019;32(2).

[27] World Health Organization. Ending the neglect to attain the sustainable development goals: A roadmap for neglected tropical diseases 2021-2030. 2020;.

[28] Deplazes P, Rinaldi L, Alvarez RC, Torgerson P, Harandi M, Romig T, et al. Global Distribution of Alveolar and Cystic Echinococcosis. Advances in Parasitology. 2017;95:315.

[29] Gripper LB, Welburn SC. The causal relationship between neurocysticercosis infection and the development of epilepsy - a systematic review. Infectious Diseases of Poverty. 2017;6(31).

[30] Rajshekhar V. Neurocysticercosis: Diagnostic problems current therapeutic strategies. Indian Journal of Medical Research. 2016;144(3):319–326.

[31] Braae UC, Devleesschauwer B, Gabriël S, Dorny P, Speybroeck N, Magnussen P, et al. CystiSim–an agent-based model for Taenia solium transmission and control. PLoS Neglected Tropical Diseases. 2016;10(12).

[32] World Health Organization. Strongyloidiasis Epidemiology;.

[33] Centers for Disease Control and Prevention. Strongyloidiasis Epidemiology and Risk Factors. 2018;.

[34] Fleitas PE, Kehl SD, Lopez W, Travacio M, Nieves E, Gil JF, et al. Mapping the global distribution of Strongyloides stercoralis and hookworms by ecological niche modeling. Parasites & Vectors. 2022;15(1):1–12.

[35] Cabada MM, Morales ML, Lopez M, Reynolds ST, Vilchez EC, Lescano AG, et al. Hymenolepis nana Impact among Children in the Highlands of Cusco, Peru: An Emerging Neglected Parasite Infection. The American Journal of Tropical Medicine and Hygiene. 2016;95(5):1031–1036.

[36] Vilchez Barreto PM, Gamboa R, Santivanez R, O’Neal SE, Muro C, Lescano AG, et al. Prevalence, Age Profile, and Associated Risk Factors for Hymenolepis nana Infection in a Large Population-Based Study in Northern Peru. The American Journal of Tropical Medicine and Hygiene. 2017;97(2):583–586.

[37] Magalhaes RJS, Fancony C, Gamboa D, Langa AJ, Sousa-Figueiredo JC, Clements AC, et al. Extending helminth control beyond STH and schistosomiasis: the case of human hymenolepiasis. PLoS Neglected Tropical Diseases. 2013;7(10):e2321.

[38] Kinyoki DK, Osgood-Zimmerman AE, Pickering BV, Schaeffer LE, Marczak LB, Lazzar-Atwood A, et al. Mapping child growth failure across low-and middle-income countries. Nature. 2020;577(7789):231–234.

[39] Cowie RH. Biology, Systematics, Life Cycle, and Distribution of Angiostrongylus cantonensis, the Cause of Rat Lungworm Disease. Hawai’i Journal of Medicine and Public Health. 2013;72(6 Suppl 2):6–9.

[40] Martins YC, Tanowitz HB, Kazacos KR. Central nervous system manifestations of Angiostrongylus cantonensis infection. Acta Tropica. 2014;141:46–53.

[41] Lu XT, Gu QY, Limpanont Y, Song LG, Wu ZD, Okanurak K, et al. Snail-borne parasitic diseases: an update on global epidemiological distribution, transmission interruption and control methods. Infectious diseases of poverty. 2018;7(1):1–16.

[42] Mediannikov O, Ranque S. Mansonellosis, the most neglected human filariasis. New Microbes and New Infections. 2018;26:S19–S22.

[43] Ta-Tang TH, Crainey JL, Post RJ, Luz SL, Rubio JM. Mansonellosis: current perspectives. Research and Reports in Tropical Medicine. 2018;9:9–24.

[44] Molyneux D, Sankara DP. Guinea worm eradication: Progress and challenges—should we beware of the dog? PLoS Neglected Tropical Diseases. 2017;11(4):e0005495.

[45] Wilson-Aggarwal JK, Goodwin CE, Swan GJ, Fielding H, Tadesse Z, Getahun D, et al. Ecology of domestic dogs (Canis familiaris) as a host for Guinea worm (Dracunculus medinensis) infection in Ethiopia. Transboundary and Emerging Diseases. 2020;.

[46] May JM. Map of the world distribution of helminthiases. Geographical Review. 1952;42(1):98– 101.

[47] Boyce MR, Carlin EP, Schermerhorn J, Standley CJ. A One Health approach for Guinea worm disease control: scope and opportunities. Tropical medicine and infectious disease. 2020;5(4):159.

[48] Sripa B, Bethony JM, Sithithaworn P, Kaewkes S, Mairiang E, Loukas A, et al. Opisthorchiasis and Opisthorchis-associated cholangiocarcinoma in Thailand and Laos. Acta Tropica. 2010;120(Supplement 1):S158–S168.

[49] Banales JM, Cardinale V, Carpino G, Marzioni M, Andersen JB, Invernizzi P, et al. Expert consensus document: Cholangiocarcinoma: current knowledge and future perspectives consensus statement from the European Network for the Study of Cholangiocarcinoma (ENS-CCA). Nature Reviews Gastroenterology and Hepatology. 2016;13(5):261–280.

[50] Kim TS, Pak JH, Kim JB, Bahk YY. Clonorchis sinensis, an oriental liver fluke, as a human biological agent of cholangiocarcinoma: a brief review. BMB Reports. 2016;49(11):590–597.

[51] Li CD, Yang HL, Wang Y. Capillaria hepatica in China. World Journal of Gastroenterology. 2010;16(6):698–702.

[52] Saichua P, Nithikathkul C, Kaewpitoon N. Human intestinal capillariasis in Thailand. World Journal of Gastroenterology. 2008;14(4):506–510.

[53] Chai JY, Shin EH, Lee SH, Rim HJ. Foodborne Intestinal Flukes in Southeast Asia. The Korean Journal of Parasitology. 2009;47:S69–S102.

[54] Mas-Coma S, Bargues M, Valero M. Gastrodiscoidiasis, a plant-borne zoonotic disease caused by the intestinal amphistome fluke Gastrodiscoides hominis (Trematoda: Gastrodiscidae). Revista Ibérica de Parasitología. 2005;66:75–81.

[55] Dada-Adegbola H, Falade C, Oluwatoba O, Abiodun O. Gastrodiscoides hominis infection in a Nigerian-case report. West African Journal of Medicine. 2004;23(2):185–186.

[56] Blum AJ, Hotez PJ. Global “worming”: Climate change and its projected general impact on human helminth infections. PLoS Neglected Tropical Diseases. 2018;12(7).

[57] Carlson CJ, Bannon E, Mendenhall E, Newfield T, Bansal S. Rapid range shifts in African Anopheles mosquitoes over the last century. bioRxiv. 2019;p. 673913.

[58] Carlson CJ, Burgio KR, Dougherty ER, Phillips AJ, Bueno VM, Clements CF, et al. Parasite biodiversity faces extinction and redistribution in a changing climate. Science Advances. 2017;3(9):e1602422.

[59] York EM, Butler CJ, Lord WD. Global decline in suitable habitat for Angiostrongylus (= Parastrongylus) cantonensis: the role of climate change. PLoS One. 2014;9(8):e103831.

